# Viral Cultures for Assessing Fomites Transmission of SARS-CoV-2: a Systematic

**DOI:** 10.1101/2022.01.26.22269917

**Authors:** Igho J. Onakpoya, Carl J. Heneghan, Elizabeth A. Spencer, Jon Brassey, Elena C. Rosca, Susanna Maltoni, Annette Plüddemann, David H. Evans, John M. Conly, Tom Jefferson

## Abstract

This is a protocol for a systematic review to assess fomite transmission in SARS-CoV-2. Our research questions are as follows:

1. Are fomite samples infectious?
2. If so, what proportion are infectious, and what is the distance and duration of infectiousness in the air?
3. What is the relationship between fomites, infectiousness and PCR cycle threshold (Ct)?
4. Is there evidence of a chain of transmission that establishes an actual instance of fomite transmission of SARS-CoV-2?

We will include studies of any design (and in any setting) that investigate fomite transmission (defined as any inanimate object that, when contaminated with or exposed to infectious agents, can transfer the agent to a new host). We will only include studies that performed viral culture which assessed cytopathic effect and verification techniques to ensure the cultured virus is SARS-CoV-2. We will assess the risk of bias using a checklist modified from the QUADAS-2 criteria.

## INTRODUCTION

The SARS-CoV-2 (COVID-19) pandemic continues to be a major public health concern. Based on WHO statistics, there have been over 265 million confirmed cases and over five million deaths globally as of 8th December 2021 [1]. Although several vaccines have been developed and programmes have been implemented globally, mutations in the virus have resulted in the occurrence of variants that reduce their effectiveness [2]. In addition, the duration of protection appears to be limited [3] and booster doses are being promoted [4].

Aside from the use of vaccines, a better understanding of the transmission dynamics of the virus will help with developing interventions that can interrupt the chain of transmission and reduce the spread of infection. We previously found that the evidence from published studies assessing the risk of transmission of SARS-CoV-2 via fomites was limited [5].

Furthermore, the quality of the included primary studies was low to very low, probably due to limited understanding of the dynamics of SARS-CoV-2 shedding at the onset of the pandemic, the types of studies done, the timing of collection, and sampling issues. In addition, less than a fifth of included studies at that time examined the cytopathic effect of SARS-CoV-2 and relied only on detecting RNA from SARS-CoV-2. Since we published that review, several studies examining the transmission of SARS-CoV-2 via fomites have been published. Therefore, it is necessary to update the evidence on fomite transmission using the highest quality evidence from such published studies.

To assess the transmission potential of fomites of SARS-CoV-2, we aim to address the following questions:

## METHODS

We aim to identify, appraise, and summarise the evidence relating to the role of fomite transmission of SARS-CoV-2 and its relationship with infectiousness (viral culture and/or serial qRT-PCRs with or without gene sequencing) and the factors influencing transmissibility.

### Search Strategy

We will conduct searches in the WHO Covid-19 Database, LitCovid, medRxiv, and Google Scholar for SARS-CoV-2 using keywords and associated synonyms. The searches for this update will be conducted up to 31st December 2021. No language restrictions will be imposed. An information specialist (JB) will conduct the searches, and for relevant papers, will undertake forward citation to identify relevant studies (see Appendix A). Two reviewers (IJO, EAS) will independently screen study abstracts to determine eligibility. Any disagreements will be resolved through discussion. Where a consensus cannot be reached, a third reviewer (TJ) will arbitrate.

### Data Extraction

We will include studies of any design (and in any setting) that investigate fomite transmission (defined as any inanimate object that, when contaminated with or exposed to infectious agents, can transfer the agent to a new host).

We will only include studies that performed viral culture which assessed cytopathic effect and verification techniques to ensure the cultured virus is SARS-CoV-2. Studies that performed serial qRT-PCR (with or without genomic sequencing) in addition to viral culture will also be included. Predictive or modelling studies will be excluded. Results will be reviewed for relevance and for articles that look relevant, forward citation matching will be undertaken to ensure relevant studies are identified.

We have previously defined viral culture as encompassing several methods that can uniquely identify the replicating agent as SARS-CoV-2. [6] Most commonly, this would be a plaque assay combined with a PCR diagnosis or immunological staining or gene sequencing of viral RNA. Viral genome sequencing is a process that helps to determine the order, or sequence, of the nucleotides in each of the genes present in a virus’s genome [see https://www.cdc.gov/flu/about/professionals/genetic-characterization.htm].

To assess the chain of transmission (question 4), we will only include studies with (a) documentation of the likelihood of transmission; (b) presence of infectious virus from viral culture (defined as encompassing any of several methods whereby one can detect exponential virus growth in cell culture in combination with a method that can uniquely identify the replicating agent as being SARS-CoV-2) and/or documentation of phylogenetics (i.e., genetic sequence lineage); and/or (c) adequate follow-up and reporting of symptoms and signs [6, 7] See supplementary material, on figshare. Dataset Available at https://doi.org/10.6084/m9.figshare.18334898

Appendix A: Search Strategy

Appendix B: Transmission Assessment Explainer

Appendix C: Standardised transmission of SARS-CoV-2 case causality assessment

We will extract the following information from included studies: study characteristics, setting, population (if any), main methods, and associated outcomes including the number of swab samples taken, the frequency and timing of samples, methods used for sample collection, hygiene practises, cycle thresholds (Cts), use of internal controls for Cts, determination of which platform was used and which genes were targeted, and sample concentrations using one or more techniques where reported. We will also extract the information on viral cultures including the methods (timing, media), verification techniques, quantification, and the final reported results. One reviewer (IJO) will extract data from the included studies, and these will be independently verified by a second reviewer (EAS). Any disagreements will be resolved through discussion.

### Quality assessment

We will assess the risk of bias modified from the QUADAS-2 criteria and previous published methods [6, 8].

We will assess the following domains:

i. Source population – was the population recruited into the study clearly described?
ii. Methods – did the study authors sufficiently describe the methods used to enable replication of the study
iii. Sample sources – were sources for the fomites samples clear?
iv. Outcome reporting – was the analysis of the results appropriate, and
v. Follow-up – was the pattern and number of fomites samples sufficient to demonstrate fomite transmission.

Where necessary, one reviewer will contact the corresponding authors of the included papers for additional information. We will also include the authors’ responses to requests for additional information in our assessment of bias. One reviewer (IJO) will categorise the potential for bias as high, moderate, or low which was independently checked by a second reviewer (EAS). Reasons for the bias assessment for each study will also be recorded. Disagreements will be resolved through discussion with the help of a third reviewer (CJH).

## Data Availability

All data included in the review and the Appendices, tables and text will be made available at Figshare

https://doi.org/10.6084/m9.figshare.18334898

## Data Analysis

We will follow PRISMA reporting guidelines as indicated for systematic or scoping reviews where applicable [9]. We will present the frequency of positive tests, and summary tables to present cycle thresholds, the results of cytopathic effects, information on viral cultures including the methods (timing, media), verification techniques, and quantification when reported. Where possible, we will report subgroup analyses by study setting, type of fomite, frequency of contacts with the fomite surface, and methods used for performing viral culture and timing for assessing cytopathic effect.

## Ethics committee approval

No ethics approval is necessary.

## Data Availability

All data included in the review and the Appendices, tables and text will be made available at Figshare: https://doi.org/10.6084/m9.figshare.18334898 files available to download.

## Funding

This work is part-funded by the NIHR School for Primary Care Research [project 569]. The work also received funding from the University of Calgary. The World Health Organization funded the first iteration of this review: Living rapid review on the modes of transmission of SARs-CoV-2 reference WHO registration No 2020/1077093. CJH, AP and EAS also receive funding support from the National Institute of Health Research School of Primary Care Research Evidence Synthesis Working Group project 390. (https://www.spcr.nihr.ac.uk/eswg).

## Conflict of interest statements

TJ received a Cochrane Methods Innovations Fund grant to develop guidance on using regulatory data in Cochrane reviews (2015 to 2018). From 2014 to 2016, he was a member of three advisory boards for Boehringer Ingelheim. TJ was a member of an independent data monitoring committee for a Sanofi Pasteur clinical trial on an influenza vaccine. Market research companies occasionally interview TJ about phase I or II pharmaceutical products for which he receives fees (current). TJ was a member of three advisory boards for Boehringer Ingelheim (2014 to 16). TJ was a member of an independent data monitoring committee for a Sanofi Pasteur clinical trial on an influenza vaccine (2015 to 2017). TJ is a relator in a False Claims Act lawsuit on behalf of the United States that involves sales of Tamiflu for pandemic stockpiling. If resolved in the United States favour, he would be entitled to a percentage of the recovery. TJ is coholder of a Laura and John Arnold Foundation grant for the development of a RIAT support centre (2017 to 2020) and Jean Monnet Network Grant, 2017 to 2020 for The Jean Monnet Health Law and Policy Network. TJ is an unpaid collaborator to the Beyond Transparency in Pharmaceutical Research and Regulation led by Dalhousie University and funded by the Canadian Institutes of Health Research (2018 to 2022). TJ consulted for Illumina LLC on next-generation gene sequencing (2019 to 2020). TJ was the consultant scientific coordinator for the HTA Medical Technology programme of the Agenzia per I Servizi Sanitari Nazionali (AGENAS) of the Italian MoH (2007 to 2019). TJ is Director Medical Affairs for BC Solutions, a market access company for medical devices in Europe. TJ was funded by NIHR UK and the World Health Organization (WHO) to update Cochrane review A122, Physical Interventions to interrupt the spread of respiratory viruses. Oxford University funds TJ to carry out a living review on the transmission epidemiology of COVID 19. Since 2020, TJ receives fees for articles published by The Spectator and other media outlets. TJ is part of a review group carrying out a Living rapid literature review on the modes of transmission of SARS CoV 2 (WHO Registration 2020/1077093 0). He is a member of the WHO COVID 19 Infection Prevention and Control Research Working Group, for which he receives no funds. TJ is funded to co-author rapid reviews on the impact of Covid restrictions by the Collateral Global Organisation.

CJH holds grant funding from the NIHR, the NIHR School of Primary Care Research, the NIHR BRC Oxford and the World Health Organization for a series of Living rapid reviews on the modes of transmission of SARs CoV 2, reference WHO registration No2020/1077093, and to carry out a scoping review of systematic reviews of interventions to improve vaccination uptake, reference WHO Registration 2021/1138353-0. He has received financial remuneration from an asbestos case and given legal advice on mesh and hormone pregnancy tests cases. He has received expenses and fees for his media work, including occasional payments from BBC Radio 4 Inside Health and The Spectator. He receives expenses for teaching EBM and is also paid for his GP work in NHS out of hours (contract Oxford Health NHS Foundation Trust). He has also received income from the publication of a series of toolkit books and appraising treatment recommendations in non-NHS settings. He is the Director of CEBM, an NIHR Senior Investigator and an advisor to Collateral Global.

DHE holds grant funding from the Canadian Institutes for Health Research and Li Ka Shing Institute of Virology relating to the development of Covid 19 vaccines and the Canadian Natural Science and Engineering Research Council concerning Covid 19 aerosol transmission. He is a recipient of World Health Organization and Province of Alberta funding which supports the provision of BSL3 based SARS CoV 2 culture services to regional investigators. He also holds public and private sector contract funding relating to the development of poxvirus based Covid 19 vaccines, SARS CoV 2 inactivation technologies, and serum neutralization testing.

JMC holds grants from the Canadian Institutes for Health Research on acute and primary care preparedness for COVID 19 in Alberta, Canada and was the primary local Investigator for a *Staphylococcus aureus* vaccine study funded by Pfizer, for which all funding was provided only to the University of Calgary. He is a co-investigator on a WHO funded study using integrated human factors and ethnography approaches to identify and scale innovative IPC guidance implementation supports in primary care with a focus on low resource settings and using drone aerial systems to deliver medical supplies and PPE to remote First Nations communities during the COVID 19 pandemic. He also received support from the Centers for Disease Control and Prevention (CDC) to attend an Infection Control Think Tank Meeting. He is a member and Chair of the WHO Infection Prevention and Control Research and Development Expert Group for COVID 19 and the WHO Health Emergencies Programme (WHE) Ad hoc COVID 19 IPC Guidance Development Group, both of which provide multidisciplinary advice to the WHO, for which no funding is received and from which no funding recommendations are made for any WHO contracts or grants. He is also a member of the Cochrane Acute Respiratory Infections Group.

JB is a major shareholder in the Trip Database search engine (www.tripdatabase.com) as well as being an employee. In relation to this work, Trip has worked with a large number of organizations over the years; none have any links with this work. The main current projects are with AXA and Collateral Global.

ECR was a member of the European Federation of Neurological Societies(EFNS) / European Academy of Neurology (EAN) Scientist Panel, Subcommittee of Infectious Diseases (2013 to 2017). Since 2021, she is a member of the International Parkinson and Movement Disorder Society (MDS) Multiple System Atrophy Study Group, the Mild Cognitive Impairment in Parkinson Disease Study Group, and the Infection Related Movement Disorders Study Group. She was an External Expert and sometimes Rapporteur for COST proposals (2013, 2016, 2017, 2018, 2019) for Neurology projects. She is a Scientific Officer for the Romanian National Council for Scientific Research.

SM is a pharmacist working for the Italian National Health System since 2002 and a member of one of the three Institutional Review Boards of Emilia-Romagna Region (Comitato Etico Area Vasta Emilia Centro) since 2018.

AP holds grant funding from the NIHR School of Primary Care Research.

IJO and EAS have no interests to disclose.

## Notes

### Funding Statement

This work is part-funded by NIHR School for Primary Care Research [project 569]. The work also received funding from the University of Calgary.

